# ClinGen *CDH1* specifications for the ACMG/AMP guidelines: improvement of germline variant clinical assertions and updated curation guidelines

**DOI:** 10.1101/2021.11.01.21265332

**Authors:** Xi Luo, Jamie L. Maciaszek, Bryony A. Thompson, Huei San Leong, Katherine Dixon, Sónia Sousa, Michael Anderson, Maegan E. Roberts, Kristy Lee, Amanda B. Spurdle, Arjen R. Mensenkamp, Terra Brannan, Carolina Pardo, Liying Zhang, Tina Pesaran, Sainan Wei, Grace-Ann Fasaye, Chimene Kesserwan, Brian H. Shirts, Jeremy L. Davis, Carla Oliveira, Sharon E. Plon, Kasmintan A. Schrader, Rachid Karam, on behalf of the ClinGen *CDH1* Variant Curation Expert Panel

**Affiliations:** Department of Pediatrics/Hematology-Oncology, Baylor College of Medicine, Houston, TX, USA; Department of Pathology, St. Jude Children’s Research Hospital, Memphis, TN, USA; Department of Pathology, Royal Melbourne Hospital, Parkville, Victoria, Australia; Department of Pathology, Peter MacCallum Cancer Centre, Parkville, VIC, Australia; Department of Medical Genetics, University of British Columbia, Vancouver, BC, CAN; Instituto de Investigac□ão e Inovac□ão em Saúde – (i3S), Institute of Molecular Pathology and Immunology of the University of Porto – (Ipatimup), Faculty of Medicine, University of Porto, Porto, PRT; Invitae Corporation, San Francisco, CA, USA; GeneDx, Gaithersburg, MD, USA; Department of Genetics, University of North Carolina at Chapel Hill, Chapel Hill, NC, USA; QIMR Berghofer Medical Research Institute, Brisbane, AUS; Department of Human Genetics, Radboud University Medical Centre, Nijmegen, NLD; Ambry Genetics, Aliso Viejo, CA, USA; Department of Pathology and Laboratory Medicine, University of California, Los Angeles, CA, USA; Department of Pathology, University of Kentucky, Lexington, KY, USA; Center for Cancer Research, National Cancer Institute, National Institutes of Health, Bethesda, Maryland USA; Department of Laboratory Medicine and Pathology, University of Washington, Seattle, WA, USA

## Abstract

**Purpose:** The Clinical Genome Resource (ClinGen) *CDH1* Variant Curation Expert Panel (VCEP) developed specifications for *CDH1* variant curation with a goal to resolve variants of uncertain significance (VUS) and with ClinVar conflicting interpretations for effective medical care. In addition, the *CDH1* VCEP continues to update these specifications in keeping with evolving clinical practice and variant interpretation guidelines.

**Methods:** *CDH1* variant classification specifications were modified based on updated genetic testing clinical criteria, new recommendations from ClinGen, and expert knowledge from ongoing *CDH1* variant curations. Trained biocurators curated 273 variants using updated *CDH1* interpretation guidelines and incorporated published and unpublished data provided by diagnostic laboratories. All variants were reviewed by the ClinGen VCEP and classifications submitted to ClinVar.

**Results:** Updated *CDH1*-specific variant interpretation guidelines include eleven major modifications since the initial specifications from 2018. Using the refined guidelines, 97% (36/37) of variants with ClinVar conflicting interpretations were resolved into benign, likely benign, likely pathogenic, or pathogenic, and 35% (15/43) of VUS were resolved into benign or likely benign. Overall, 88% (239/273) of curated variants had non-VUS classifications.

**Conclusion:** The development and evolution of *CDH1-*specific criteria by the expert panel results in decreased uncertain and conflicting interpretations of variants in this clinically actionable gene.

## 1. Introduction

The National Institutes of Health (NIH) developed the Clinical Genome Resource (ClinGen) to standardize the clinical annotation and interpretation of genomic variants and to implement evidence-based expert consensus for curating genes and variants^1^. To achieve these goals, Variant Curation Expert Panels (VCEPs) are formed in the ClinGen program with a focus on specific genes and diseases. VCEPs develop gene- or disease-specific variant classification rules based on the American College of Medical Genetics and Genomics and the Association for Molecular Pathology (ACMG/AMP) classification framework and curate genetic variants to be deposited in ClinVar^2^ (https://www.ncbi.nlm.nih.gov/clinvar/). In ClinVar, VCEP-interpretations receive a three-star review level status (reviewed by expert panel)^1^ and designation that the interpretation process is recognized by the U.S. Food and Drug Administration (FDA)^3,4^.

Germline pathogenic (P) or likely pathogenic (LP) variants in the *CDH1* gene (NM_004360.5) predispose to hereditary diffuse gastric cancer (HDGC; MIM# 137215), a cancer susceptibility syndrome inherited in an autosomal dominant pattern, characterized by increased risk for diffuse gastric cancer (DGC) as well as lobular breast cancer (LBC). Risk-reduction strategies for heterozygotes of P and LP *CDH1* variants include consideration for total gastrectomy. Female heterozygotes may also consider bilateral mastectomy^5^. The ClinGen Hereditary Cancer Clinical Domain Working Group selected *CDH1* as a high priority gene and convened a *CDH1* VCEP in 2015. The initial *CDH1*-specific variant interpretation guidelines were published in 2018^6^. These guidelines apply only to HDGC and not to other *CDH1*-associated conditions, such as blepharocheilodontic syndrome (MIM# 119580).

Over the past three years, there has become a clear need to update the *CDH1* specifications based on: learned experiences from ongoing *CDH1* variant curations; updated HDGC clinical practice guidelines which expanded *CDH1* genetic testing recommendations^5^; and general recommendations for the refinement of the ACMG/AMP criteria provided by the ClinGen Sequence Variant Interpretation (SVI) working group. At this time, there have been two updates to our interpretation guidelines which are both described in this paper: version 2 (published 9/6/2019) and version 3 (published 9/20/2021). At any time, the most recent guidelines can be found at: https://www.clinicalgenome.org/affiliation/50014/. Modifications to the ClinGen *CDH1* variant interpretation guidelines illustrate the dynamic evolution of ClinGen expert panel specifications that incorporate up-to-date knowledge and inform evidence-based interpretations to support and benefit precision medicine and clinical research.

Since publication of the initial interpretation guidelines, the *CDH1* VCEP has curated 273 *CDH1* variants and deposited these interpretations into the ClinVar database^2^. Importantly, only 12% (34) of these submissions are variants of uncertain significance (VUS) after curation using updated *CDH1* variant interpretation guidelines. The resolution of both VUS and variants with conflicting interpretations from ClinVar submitters is critical to provide diagnostic information to clinicians and patients and allow for informed patient care.

## 2. Methods

### 2.1 ClinGen *CDH1* VCEP and updates to variant interpretation guidelines

The *CDH1* VCEP comprises 32 professionals with expertise in key domains regarding HDGC and/or variant classification and includes clinicians, research scientists, genetic counselors, pathologists, and clinical laboratory diagnosticians. Currently, there are representatives from 22 participating institutions in six countries: Australia, Canada, Italy, Netherlands, Portugal, and the United States. The *CDH1* VCEP also includes representatives from several diagnostic laboratories performing *CDH1* germline testing and research.

Modifications to the *CDH1* variant interpretation guidelines were proposed and discussed via teleconference calls and e-mail communications to arrive at consensus decisions, followed by submission to and approval by the ClinGen Sequence Variant Interpretation working group (SVI). At the time of publication, the VCEP has submitted two updates to the ClinGen SVI for approval (https://www.clinicalgenome.org/affiliation/50014/). Version 2 and Version 3 of the guidelines were published to the ClinGen website on 9/6/2019 and 9/20/2021, respectively. The current work incorporates both updates.

### 2.2 Curation process and variant classification

*CDH1* variants were curated and classified according to the current *CDH1*-specific variant interpretation guidelines at the time of curation. Reclassification using the version 3 guidelines was performed for all variants. Variants prioritized for expert panel review included: 1) variants with conflicting interpretations (CI) in ClinVar; 2) nonsense, frameshift, initiation codon, and canonical splice site variants; 3) variants with an allele frequency of ≥ 0.2% in population databases; 4) VUS with a two-star review status in ClinVar; and 5) internal or external requests for expert interpretation.

Trained ClinGen biocurators utilized ClinGen’s Variant Curation Interface (https://curation.clinicalgenome.org/) to aggregate and assess data and document the applicable criteria for each variant. In addition to published case evidence from literature, unpublished and de-identified patient observations were provided by VCEP members, laboratory representatives, and the NCI hereditary gastric cancer study (Clinicaltrials.gov NCT03030404) to facilitate comprehensive variant interpretation. Biocurators assigned a provisional classification of benign (B), likely benign (LB), VUS, LP, or P to each variant. Variants were reviewed by expert members during monthly *CDH1* VCEP meetings to modify and approve the classification of variants. The final variant classifications were submitted to ClinVar with three-star level review status and FDA-recognized designation (https://www.ncbi.nlm.nih.gov/clinvar/submitters/506817/).

## 3. Results

### 3.1 Updates to ClinGen *CDH1* VCEP specifications to the ACMG/AMP variant interpretation guidelines

Since the initial *CDH1*-specifications to the ACMG/AMP variant classification guidelines were published in 2018^6^, we have made 11 major updates to these criteria which are summarized here and described below and in Table 1:

1. Adoption of genetic testing criteria used to define HDGC phenotype criteria from the 2020 updated HDGC clinical practice guidelines^5^ for PS2, PS4, PM6, and PP1 (v3);
2. Application of PVS1 to initiation codon variants (v2);
3. Development of a decision tree to specify *CDH1* PVS1 rules (v3) (figure 1);
4. Integration of splice site specific recommendations to determine the strength of PVS1 criterion (v3);
5. Approval of BS1 or BS2 alone as adequate criteria for LB classification (v2);
6. Specification of PM5_supporting to nonsense and frameshift variants that are predicted and/or proven to undergo nonsense-mediated decay (NMD), or to splicing variants in acceptor/donor sites that have at least one other variant meeting P/LP at the same site (v3);
7. Downgrade of PM2 to supporting strength (v3);
8. Removal of PS1 as an applicable evidence code (v3);
9. Removal of the conservation requirement from BP7 and expansion of BP7 to intronic variants at or within +7 to -21 locations (v3);
10. Use of the Bayesian point system^7,8^ in curations with conflicting evidence that otherwise result in a classification of VUS (v3);
11. Clarification on the usage of PS4, PM2, PM4, PP1, BS2, BS3, BP2, BP4, and BP5 (v2 and v3).

**Figure 1:**
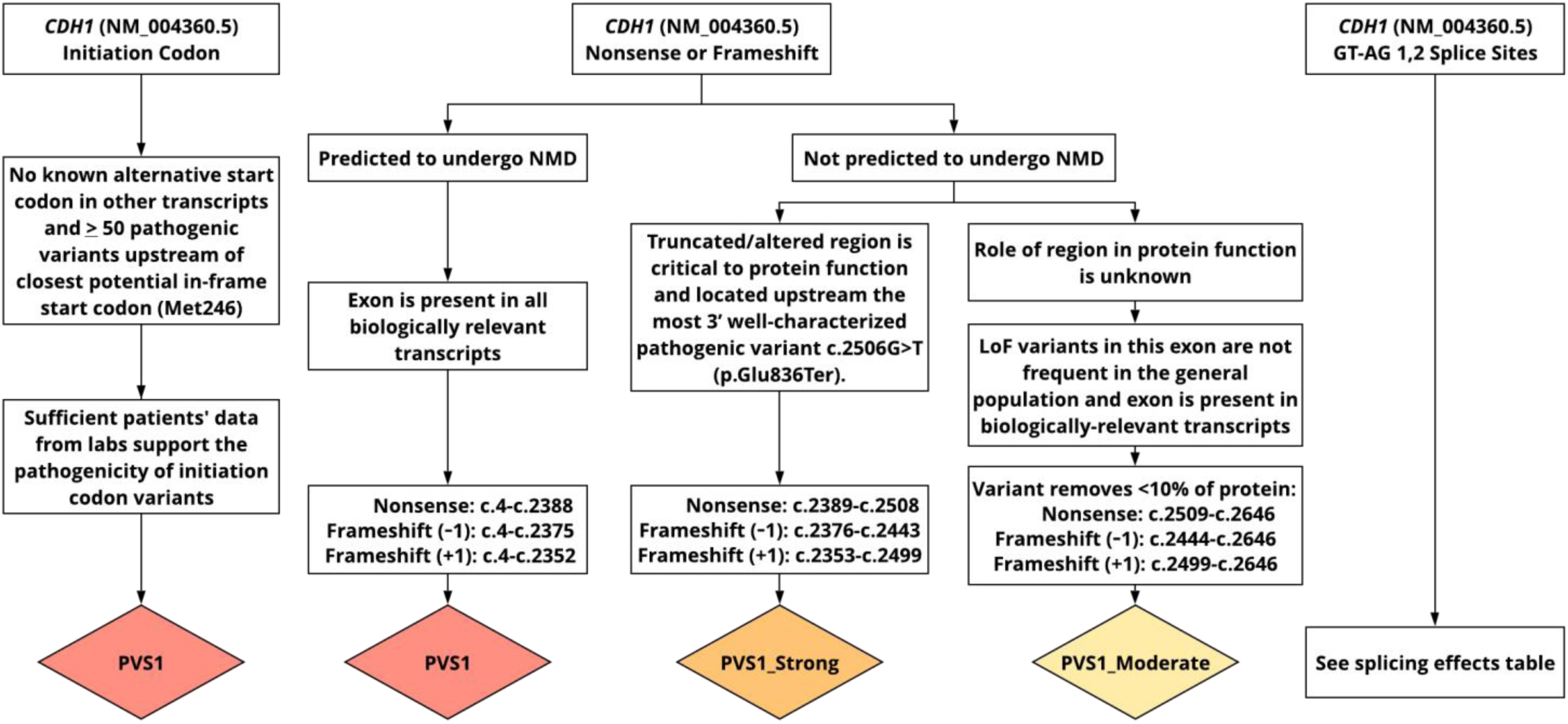
PVS1 decision tree for *CDH1*. A gene-specific decision tree was developed to clarify rules for application of PVS1 (adapted from Tayoun et al 2018^13^). Initiation codon variants and nonsense or frameshift variants predicted to undergo NMD should justify the PVS1 criterion at very strong strength. For truncations in the NMD-resistant zone, PVS1 should be applied at strong or moderate strength depending on whether the location of the variant is upstream or downstream of the most 3’ well-characterized pathogenic variant [NM_004360.5:c.2506G>T (p.Glu836Ter)]. Refer to Supplementary Table S3 for key splice site information including the recommended strength of the PVS1 criterion for each donor and acceptor site.

### Three minor updates were also made to the *CDH1* variant curation criteria

1. Updating PP3 and BP4 to include SpliceAI^9^ as a recommended splicing predictor (v3);
2. Specifying a subpopulation minimum threshold of 2,000 alleles for the application of BA1 and BS1^10^ (v3);
3. Specifying a mean coverage of at least 30X coverage from next generation sequencing data for application of BA1, BS1, and PM2^11^ (v3);

Ongoing updates will allow for continued improvement of *CDH1* variant curations resulting in comprehensive and confident classifications.

#### 3.1.1 Implementation of updated HDGC clinical practice guidelines for specification of *de novo* (PS2, PM6), phenotype (PS4), and segregation (PP1) data

The updated International Gastric Linkage Consortium 2020 HDGC genetic testing criteria^5^ have expanded the recommendation for *CDH1* germline testing for individuals with isolated DGC to those who are younger than 50 years of age. To apply patient-related criteria, including *de novo* (PS2, PM6), phenotype (PS4), and familial segregation (PP1), all probands being counted should have a personal and/or family history meeting one of the HDGC criteria in Supplementary Table S1. Individuals in the same family should only be counted towards PP1 criterion and not as additional cases to count for PS4.

For *CDH1* variant curation, application of PS4 is determined by proband counting, or the number of unrelated individuals meeting clinical criteria for HDGC. The application of PS4 in the context of *CDH1* variants should also take into consideration the fact that about 30% of the reported individuals with pathogenic variants meet the HDGC phenotypic criteria based on penetrance estimates^12^. For example, if the variant is observed in 20 families, at least six of these (30%) must meet HDGC phenotypic criteria to apply PS4. In keeping with the initial *CDH1* specifications of the ACMG/AMP guidelines^6^, the strength of PS4 increases based on the number of families meeting HDGC criteria (Table 1).

#### 3.1.2 Application of PVS1 to initiation codon variants, and a decision tree and splicing table to specify the strength of PVS1 for potential loss-of-function variants in *CDH1*

The ClinGen SVI provided a general recommendation in 2018 that PVS1 or PVS1_strong criteria should not be applied to start loss variants^13^ based on functional studies demonstrating that translational re-initiation can occur at alternative ATG or non-ATG sites downstream or upstream of the original start site^14-20^. The *CDH1* VCEP has now determined that PVS1 should be applied at very strong strength for initiation codon variants (Figure 1) based on the following: 1) there are no known *CDH1* transcripts with alternative start codons, 2) there are at least 50 P/LP classified variants upstream of the closest potential in-frame start codon at Met246, and 3) patient data (currently ten probands/families meeting HDGC criteria) support the pathogenicity of initiation codon variants (Supplementary Table S2). By applying PVS1 to initiation codon variants, we have re-curated five variants as pathogenic (Supplementary Table S2). With PVS1_moderate applied based on the initial *CDH1* guidelines, these variants would have been classified as either LP (one) or VUS (four), demonstrating the importance of continued modification of VCEP specifications to improve variant classification and clinical reporting.

A *CDH1*-specific decision tree for PVS1 (Figure 1, adapted from Tayoun et al^13^) was developed to clarify rules for PVS1 application and the corresponding evidence strength. For canonical splicing variants, the *CDH1* VCEP also provided splice site-specific recommendations for the strength of PVS1 criteria and the application of PM5_supporting (see section 3.1.4; Supplementary Table S3). This splicing table contains key information for each splice site including the predicted or experimentally demonstrated splicing impacts and splice site variants that have been classified to date. Twenty-nine of the 35 splice site variants curated to date are P/LP, while six variants predicted or confirmed to produce in-frame transcripts remain classified as VUS, demonstrating the importance of splice site-specific recommendations for the interpretation of *CDH1* splicing variants.

#### 3.1.3 BS1 and BS2 are sufficient criteria for likely benign classification

For a variant to reach a LB classification, the 2015 ACMG/AMP rules require either one strong and one supporting criteria, or at least two supporting criteria. Based on this, variants meeting either BS1 (allele frequency is greater than expected for disorder) or BS2 (observed in a healthy adult individuals) alone would remain as VUS. The recent Bayesian formulation of the ACMG/AMP guidelines^7,8^, further described in section 3.1.8, indicated that strong benign evidence criteria hold a stronger weight (equal to -4 points) than two supporting benign criteria (equal to -2), with the minimum points needed for a LB classification being -1^8^. For non-synonymous variants that do not affect splicing, only two benign supporting codes are applicable, *cis*/*trans* testing (BP2) and alternate locus observations (BP5), neither of which is readily available from laboratories. Therefore, we recommended that variants meeting either BS1 or BS2 be classified as LB even in the absence of additional supporting evidence since this equates to -4 points.

Among the 24 variants classified as LB based on BS2 alone, seven were identified in ≥ 100 individuals without HDGC phenotypes (Table 2). The NM_004360.5:c.670C>T (p.Arg224Cys) variant was found in 392 unaffected individuals by clinical testing. This variant was previously curated as a VUS by the *CDH1* VCEP^6^ despite having mostly B/LB interpretations in ClinVar; the updated BS2 rule allows for consistency between the VCEP and clinical laboratories. All 24 variants classified as LB based on BS2 alone affected the ClinVar designated clinical significance of the variant (VUS/CI to LB, Table 2). In addition, seven of eight variants meeting criteria for BS1 also met criteria for BS2, with one also meeting BP4, allowing these variants to reach a benign classification. The significant improvements over ClinVar variant interpretations with the application of BS2 criterion also demonstrate the benefit of sharing unpublished, de-identified patient data from multiple diagnostic and research labs in the *CDH1* VCEP.

For application of BS2 as strong evidence, the variant must be seen in at least ten adults without a known personal and/or family history of gastric cancer, DGC, LBC, or gastric tumors with signet ring cell histology, as described in Lee et al^6^. In addition to excluding individuals with non-DGC pathology (i.e., histologically confirmed gastric cancer) and non-LBC pathology from proband counts, individuals reporting a personal and/or family history of gastric cancer without supporting pathology information should also be excluded. However, individuals with non-LBC breast cancer diagnoses should be noted for review by experts^12^. Based on penetrance estimates described in section 3.2.1^12^, BS2 cannot be applied to variants in which more than 30% of reported individuals meet HDGC criteria. As such, BS2 may not be applied in conjunction with PS4.

#### 3.1.4 PM5_supporting applies to nonsense and frameshift variants predicted to undergo NMD, and GT-AG 1,2 acceptor/donor site with P/LP splice site variants curated

The 2015 ACMG/AMP guidelines described the application of PM5 for novel missense variants occurring at the same position as another pathogenic missense change^21^. The impact of amino acid-level changes of *CDH1* variants is inconclusive, with all known P/LP *CDH1* missense variants affecting splicing (Supplementary Table S4). Therefore, we did not recommend applying PM5 to *CDH1* missense variants^6^.

After curating 113 nonsense and frameshift variants, the *CDH1* VCEP noticed a discrepancy between classifications based on gene-specific guidelines and laboratory classifications, consistent with observations from other ClinGen VCEPs. Using *CDH1* version 1 and 2 guidelines, many nonsense and frameshift variants did not have sufficient patient data to apply the PS4_supporting criterion, remaining as LP based on PVS1 and PM2. In contrast, these variants were classified as P by diagnostic laboratories regardless of the number of individuals meeting HDGC criteria. To resolve this discordance, we validated PM5_supporting for variants in exons with other confirmed loss-of-function variants, following the recommendation from the ClinGen SVI (unpublished). To validate the application of PM5_supporting to loss-of-function variants, we found that at least one pathogenic variant in each exon of *CDH1* is supported by phenotypic evidence (PS4 and/or PP1; Supplementary Table S5), indicating no exon-specific effect with respect to patient presentation, e.g., that pathogenic variants do not localize to specific exons. Exon-specific variability has been demonstrated for both *BRCA1* and *BRCA2*, where loss-of-function variants in select exons retain some of their tumor suppressor function^22^ or induce in-frame skipping through the modification of splice sites or regulatory elements resulting in a partially functional protein^23^. Based on guidance for *BRCA1* and *BRCA2*^23^, application of PM5_supporting also requires that the variant does not impact splicing based on either RNA studies or splicing predictors to ensure that the loss-of-function effect is not rescued by alternative splicing mechanisms or regulatory element modification.

Utilization of PM5_supporting has allowed 45 nonsense and frameshift variants to reach a pathogenic classification using version 3 guidelines. Implementation of this evidence code will facilitate future batch curation efforts for novel nonsense/frameshift variants in *CDH1*, reduce requests to labs and academic groups for patient phenotypic data to aid curation, and eliminate the need for re-curation of likely pathogenic variants which is required every two years.

#### 3.1.5 Decreased weight of PM2 to a supporting strength level

Consistent with guidance from the ClinGen SVI, we decreased the weight of PM2 (absence or rarity in population databases) from moderate to supporting in our version 3 guidelines. It was determined that rarity in population databases was given too much weight in the initial 2015 ACMG/AMP framework^21^ and did not meet the relative odds of pathogenicity for moderate strength evidence^7^. Recent work from the Genome Aggregation database (gnomAD) indicates that the current sample size does not capture complete mutational saturation of the human exome^24^. Similarly, findings from the ExAC database indicate that 54% of the high-quality variants are only present once in the dataset^25^. Finally, PM2 was a conflicting criteria in eight B or LB *CDH1* classifications using versions 1 and 2, supporting the strength reduction in version 3.

#### 3.1.6 Removal of PS1 as an evidence criterion

At the time of publication of the original *CDH1* specifications to the ACMG/AMP variant curation guidelines^6^, the VCEP did not feel that PS1 needed further specification.

However, evidence criteria for other codes generally applied to missense variants (PS3/BS3, PM5, PP2, and PP3/BP4) were not validated due to the paucity of missense variants classified as P/LP, and the corresponding lack of individuals with missense variants meeting diagnostic testing criteria for HDGC^6^. To date, we have curated 66 missense variants: 20 B, 22 LB, 19 VUS, 3 LP, and 2 P. All five missense variants that meet a P/LP classification affect splicing, indicating that the impact of amino acid level changes of *CDH1* variants on HDGC is inconclusive (Supplementary Table S4). The PS1 code would not be applicable for identical amino acid changes at these residues since this evidence criterion requires that the variant does not impact splicing. Based on this data, the *CDH1* VCEP has removed PS1 as an applicable evidence code.

#### 3.1.7 BP7: Removal of the requirement for nucleotide conservation and extension of evidence application to synonymous and intronic variants at or beyond +7 to - 21 locations

BP7 was originally designed to be applied to synonymous variants for which splicing prediction algorithms indicate no impact and the nucleotide is not conserved. Recent work indicates that nucleotide conservation has limited predictive power since many intronic SNVs are located at sites that are under less constraint than exonic SNVs^26^. We have therefore opted to remove the original conservation requirement. Furthermore, application of BP7 was expanded to intronic variants at or beyond +7 to -21 locations^27,28^.

#### 3.1.8 Utilization of Bayesian point system in VUS curations with conflicting evidence

For variants with conflicting pathogenic and benign criteria, the *CDH1* VCEP advises using the Bayesian point system to calculate the ultimate pathogenicity^7,8^. Specifically, we recommend utilizing the point system to resolve conflicting evidence that results in an assertion of VUS in *CDH1* curations. In 2018, Tavtigian et al. demonstrated that the ACMG/AMP guidelines are compatible with a quantitative Bayesian formulation to determine pathogenicity using four strength levels of evidence^7^. Recently, Tavtigian et al. established a natural conversion from the Bayesian formulation into a point-based system to derive variant pathogenicity^8^. Here, each evidence strength is assigned a point value where very strong evidence codes are +8 or -8 points for pathogenic or benign, respectively, strong are ±4, moderate are ±2, and supporting evidence are ±1. The summation of these point values equates to the variant classification category where ≤ -7 points corresponds to a B classification, -1 to -6 to LB, 0 to 5 to VUS, 6 to 9 to LP, and ≥ 10 to P.

### 3.2 Expert curations completed to date

As of May 20, 2021, the *CDH1* VCEP had completed the curation, expert review, and classification of 273 variants using the version 3 guidelines. All completed classifications have been submitted to ClinVar with a description of the corresponding evidence and final interpretation. The variant classifications comprise 122 P, 28 LP, 38 LB, and 51 B, while 34 (12%) classifications remain as VUS (Figure 3a).

#### 3.2.1 Comparison of *CDH1* VCEP variant classifications to previous ClinVar assertions

The ClinVar assertions of 33% (90/273) of *CDH1* VCEP variant classifications were altered using version 3 of the *CDH1* specifications. Of these, 52 (58%) changed the ClinVar designated clinical significance of the variant (VUS/CI changing to B/LB or P/LP). The remaining 38 (42%) were defined as resolution of a confidence conflict, such that variants previously classified as LB (or LP) were downgraded to B (or upgraded to P) (Figure 2).

**Figure 2:**
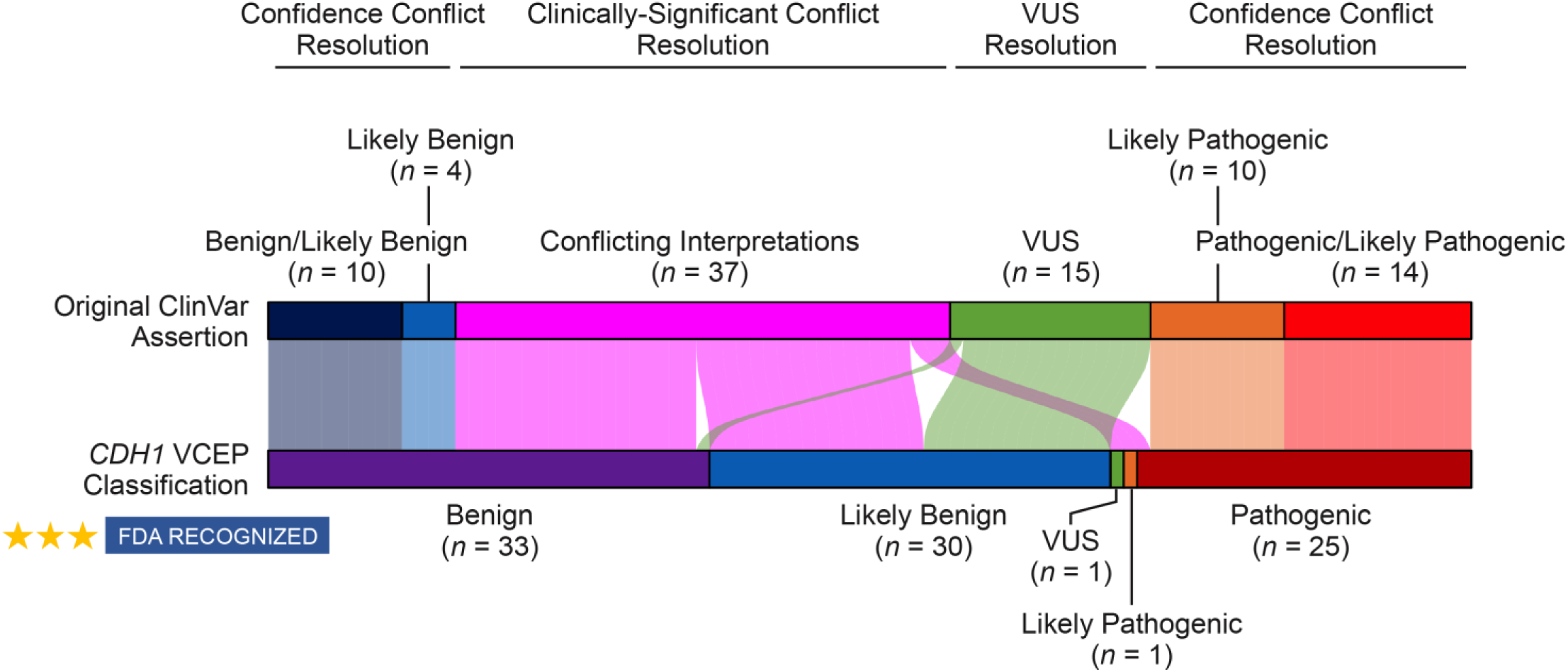
Comparison of *CDH1* VCEP variant classifications to previous ClinVar assertions. Using the *CDH1* specifications to the ACMG/AMP guidelines, 33% (90/273) variant classifications were altered and the change depicted here. Our data are limited in that ClinVar assertions for each variant were not extracted at the time of the curation and final expert interpretation of each *CDH1* variant. Therefore, the number of improved *CDH1* variant curations may be higher than reported therein. Despite this limitation in our analysis, *CDH1* expert curations still include resolution of 97% (36/37) variants with conflicting interpretations to a non-VUS classification. Eighteen variants with conflicting interpretations were resolved to benign, 16 to likely benign, one to VUS, one to likely pathogenic, and one to pathogenic. Fourteen VUS were resolved to likely benign and 1 to benign. Ten variants with a combination of benign/likely benign assertions and four variants like a likely benign assertion were resolved to benign, and 14 variants with a combination of pathogenic/likely pathogenic assertions and ten variants with a likely pathogenic assertion were resolved to pathogenic. Overall, 52 improvements resulted in resolution of a clinically significant conflict and 38 improvements resulted in resolution of a confidence conflict.

Most variants (32) with previous CI in ClinVar comprised a combination of B/LB and VUS classifications. However, five of these variants had at least one assertion of P or LP, making the classification of the variant difficult to infer based on ClinVar information. The *CDH1* VCEP reviewed variant interpretations have resolved 36 out of 37 (97%) of variants with CI to non-VUS classifications, with most of them (34) having been reclassified as B or LB (Figure 2). Notably, two variants with CI, NM_004360.5:c.1679C>G (p.Thr560Arg) and NM_004360.5:c.1057G>A (p.Glu353Lys), were reclassified as P and LP, respectively. Both are missense changes are predicted to impact splicing, with RNA assays demonstrating abnormal out-of-frame transcripts (Supplementary Table S4)^29,30^. A combination of literature reports and unpublished laboratory data indicated that these variants co-segregated in several families meeting HDGC criteria^29,31,32^. In addition, we were able to downgrade 14 VUS to LB and one to B (Figure 2, Table 2). Resolution of *CDH1* variants designated as VUS or with CI will diminish clinical uncertainty for individuals with these variants.

#### 3.2.2 Curations using each classification category

The version 3 *CDH1* variant curation guidelines utilize 19 evidence criteria established in the 2015 ACMG/AMP guidelines (Table 1). Currently, nine evidence criteria are not applicable to *CDH1*: PS1, PM1, PM3, PP2, PP4, PP5, BP1, BP3, and BP6.

With respect to *CDH1* VCEP curations reported herein (Figure 3a), the most frequently applied pathogenic evidence codes were PVS1, PM2, PS4, and PM5 (Figure 3b). Importantly, 65% of P/LP variants with PVS1 criterion applied also had supporting clinical data to meet PS4 criterion at varied strengths, indicating the importance of data sharing from laboratories. Of the 148 variants classified as P or LP, only five variants did not meet PVS1 criterion, all of which are missense variants that affect splicing (Supplementary Table S3, Supplementary Table S4), again highlighting the lack of missense variants among disease-associated variants. All variants meeting PVS1 criterion but not reaching a P or LP classification had PVS1_moderate evidence strength based on the creation of a premature stop codon downstream of NM_004360.5:c.2506G>T (p.Glu836Ter) (Five variants; Supplementary Table S5), or canonical splice sites predicted or experimentally demonstrated to result in in-frame partial exon skipping/insertion (Four variants; Supplementary Table S3, Supplementary Table S5).

**Figure 3:**
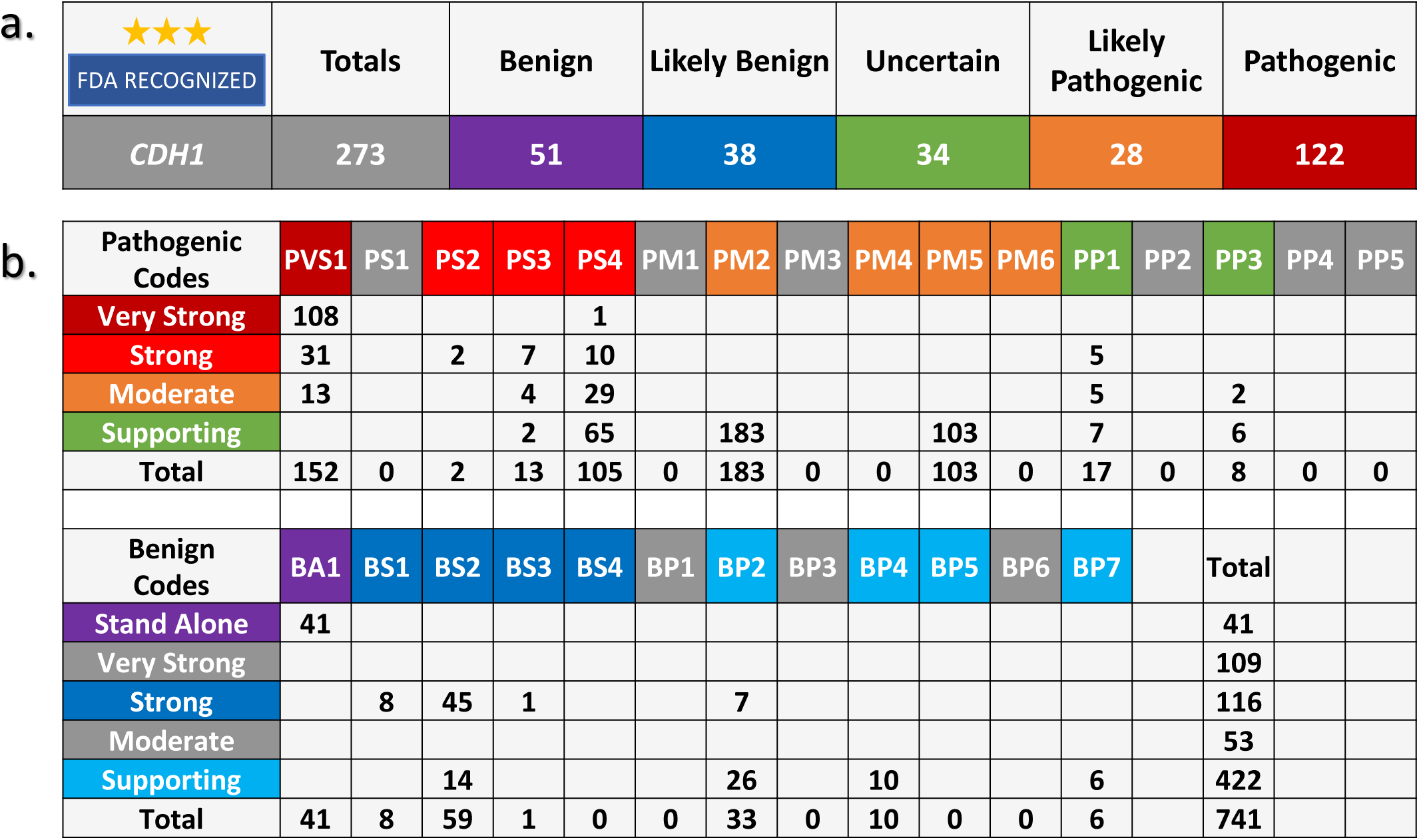
Summary of 273 *CDH1* VCEP variant interpretations and frequency of application of each evidence code. (a) The *CDH1* VCEP has curated and deposited 273 variant interpretations to the ClinVar database: 51 benign, 38 likely benign, 34 variants of uncertain significance, 28 likely pathogenic, and 122 pathogenic. (b) Frequency of application of each evidence code and its accompanying strength. The most frequently applied non-variant specific evidence codes applied were PS4, PM2, PP1, BS2, and BP2, demonstrating the importance of collating published and unpublished lab data provided by diagnostic laboratories for variant classification.

The most frequently applied benign evidence criteria were BA1, BS2, and BP2. Forty-one variants were classified as benign based on the BA1 criterion, indicating the importance of establishing gene-specific allele frequency thresholds from population databases. With the benefit of sharing internal data from multiple diagnostic and research laboratories, BS2 and BP2_strong criteria were applied to 50 variants classified as B or LB. In the absence of other evidence, application of BS2 alone allowed 24 variants to reach a LB classification, as described in section 3.1.3 (Table 2).

## Discussion

The current work highlights the critical need for gene-/disease-specific and continuously evolving ACMP/AMP guidelines for improved variant interpretation. The *CDH1* VCEP has submitted 273 expert interpretations to ClinVar. Importantly, 33% of classifications (90/273) submitted by us had improved resolution of significance over previous ClinVar classifications. These included 35% of VUS (15/43) that were resolved to B/LB and 97% of variants (36/37) with CI that were resolved into B/LB/LP/P. Overall, 88% of variants (239/273) curated by the VCEP have non-VUS classifications, whereas for *CDH1* variants not yet curated by the VCEP, only 47% have non-VUS or non-CI (as of 6/25/2021). Resolution of *CDH1* VUS and CI variants is crucial for individuals with HDGC, since only individuals with a P or LP variant in *CDH1* are recommended to undergo a risk reducing total gastrectomy.

At any time, the most up-to-date recommendations of *CDH1* evidence criteria can be found on the *CDH1* VCEP webpage (https://www.clinicalgenome.org/affiliation/50014). These variant interpretation guidelines are updated based on general recommendations from the ClinGen SVI, practical experience obtained from expert curations, and emerging evidence in specific gene and disease fields. Variant curations are ongoing and per ClinGen policy, VUS and LP variants need to be reassessed by VCEPs every two years. In addition, LB variants are required to be re-curated when a major updated population database is available (e.g., gnomAD V4), and other variants may be re-evaluated if discrepancies in the variant classification or relevant evidence becomes available. The *CDH1* VCEP will continue to submit three-star expert panel reviewed and FDA-recognized variant interpretations to ClinVar with the primary goal of resolving conflicting interpretations and VUS to ultimately improve clinical management including prophylactic surgery and cancer surveillance for HDGC patients.

## Data Availability

All variants have been submitted to ClinVar by the CDH1 VCEP (https://www.ncbi.nlm.nih.gov/clinvar/submitters/506817/) and all evidence codes utilized in variant classifications are available in the ClinGen Evidence Repository (https://erepo.clinicalgenome.org/evrepo/).

https://erepo.clinicalgenome.org/evrepo/

## Data availability

All variants have been submitted to ClinVar by the *CDH1* VCEP (https://www.ncbi.nlm.nih.gov/clinvar/submitters/506817/) and all evidence codes utilized in variant classifications are available in the ClinGen Evidence Repository (https://erepo.clinicalgenome.org/evrepo/).

## Acknowledgements

The *CDH1* VCEP thanks the ClinGen SVI Working Group as well as the Executive Committee of the Hereditary Cancer Clinical Domain Working Group.

The NIH, National Human Genome Research Institute supported this work through U41HG009649 (X.L. and S.E.P.). ABS was supported by an Australian NHMRC Investigator Fellowship (APP177524).

The ClinGen *CDH1* VCEP members include co-authors (RK and KAS as co-chairs; XL, JLM, BAT, HSL, KD, SS, MA, MER, KL, ABS, AM, TB, CP, LZ, TP, SW, GAF, CK, BHS, JLD, CO and SEP), and others, as follows: Pardeep Kaurah (BC Cancer Research Centre, CAN), Fatima Carneiro (University of Porto, PRT), Elizabeth Chao (University of California Irvine, USA), Giovanni Corso (University of Milano, ITA), Joana Figueiredo (University of Porto, PRT), David Huntsman (University of British Columbia, CAN), Sean Tavtigian (University of Utah, USA), Chella van der Post (Radboud University Medical Centre, NLD).

## Author Information

Conceptualization: XL, JLM, KAS, RK; Data curation: XL, JLM, BAT, HSL, KD, SS, SW; Formal Analysis: XL, JLM, ABS; Funding acquisition: XL, SEP; Writing – original draft: XL, JLM; Writing – review & editing: XL, JLM, BAT, HSL, KD, SS, MA, MER, KL, ABS, AM, TB, CP, LZ, TP, GAF, CK, BHS, JLD, CO, SEP, KAS, RK.

## Ethics Declaration

Not applicable

## Notes

**Conflict of Interest** The following authors are an employee, trainee or consultant for a commercial laboratory that offers genetic testing for CDH1: M.A., M.E.R., A.R.M., T.B., C.P., T.P., C.O., S.E.P. and R.K. L.Z. reports that family members hold leadership positions and ownership interests of Decipher Medicine.

### Competing Interest Statement

The following authors are an employee, trainee or consultant for a commercial laboratory that offers genetic testing for CDH1: M.A., M.E.R., A.R.M., T.B., C.P., T.P., C.O., S.E.P. and R.K. L.Z. reports that family members hold leadership positions and ownership interests of Decipher Medicine.

