## Supplementary material for "ClinGen *CDH1* specifications for the ACMG/AMP guidelines: improvement of germline variant clinical assertions and updated curation guidelines": Tables 1-2, Supplemental Tables 1-5

**Table 1: Updated *CDH1* rule specifications for the ACMG/AMP variant interpretation guidelines**

The bold text in the *CDH1* specification table indicates the updates in version 2 and 3 from the initial version of guidelines.

| ACMG/AMP Criteria Codes | Original ACMG/AMP Rule Summary | CDH1 Rule Specifications | | | | | |
| --- | --- | --- | --- | --- | --- | --- | --- |
|  |  | **Stand Alone** | **Very Strong** | **Strong** | **Moderate** | **Supporting** | **Comments** |
| PVS1 | Null variant in a gene where LoF is a known mechanism of disease | --- | **Per modified *CDH1* PVS1 decision tree** | **Per modified *CDH1* PVS1 decision tree**  Other *CDH1* caveats:  - Use **PVS1_Strong as the default** strength of evidence for canonical splice site variants **and follow the site-specific recommendations in the splicing table.**  *- CDH1* Exonic deletions or tandem duplications of in-frame exons **(exon 4,5,8,9,12,13,15)** | **Per modified *CDH1* PVS1 decision tree**  Other *CDH1* caveats:  - G to non-G variants disrupting the last nucleotide of an exon  - Canonical splice sites **predicted or** demonstrated experimentally to result in in-frame partial skipping/insertion (e.g., Exon 3 donor site) | **N/A** | RNA analysis is recommended for predicted splicing alterations, and if the RNA evidence does not support the prediction, the strength should be updated |
| PS1 | Same amino acid change as a previously established pathogenic variant regardless of nucleotide change | --- | --- | **---** | --- | --- | **Not applicable for *CDH1*** |
| PS2 | *De novo* (both maternity and paternity confirmed) in a patient with the disease and no family history | --- | ≥Two patients **meet the HDGC individual phenotype criteria** w/ parental confirmation | One patient **meets the HDGC individual phenotype criteria** w/ parental confirmation | --- | --- | Use ClinGen’s *de novo* point system for a highly specific phenotype |
| PS3 | Well-established *in vitro* or *in vivo* functional studies supportive of a damaging effect on the gene or gene product | --- | --- | RNA assay demonstrating abnormal out-of-frame transcripts | --- | RNA assay demonstrating abnormal in-frame transcripts | This rule can only be applied to demonstrate splicing defects. |
| PS4 | Prevalence of variant in affected individuals is significantly increased compared to controls | --- | **≥**Sixteen families meet HDGC criteria | **Four - Fifteen** families meet HDGC criteria | Two **or three** families meet HDGC criteria | One family meets HDGC criteria | **Use the 2020 updated clinical practice guidelines (PMID: 32758476) as the HDGC phenotype criteria.**  PS4 cannot be applied to variants that meet BS1 or BA1**, or to variants in which less than 30% of reported individuals meet HDGC criteria.** |
| PM1 | Located in a mutational hot spot and/or critical and well-established functional domain without benign variation | --- | --- | --- | --- | --- | **Not applicable for *CDH1*** |
| PM2 | Absent in population databases | --- | --- | --- | --- | **≤ One out of 100,000 alleles in gnomAD cohort; if present in ≥2 individuals within a subpopulation, must be present in ≤ One out of 50,000 alleles** | Use gnomAD to determine allele frequency. **The mean coverage of *CDH1* in the population database used should be at least 30x.** |
| PM3 | For recessive disorders, detected in trans with a pathogenic variant | --- | --- | --- | --- | --- | **Not applicable for *CDH1*** |
| PM4 | Protein length changes as a result of in-frame deletions/insertions in a nonrepeat region or stop-loss variants | --- | --- | --- | **Only apply to stop-loss variants**  Variant example: *CDH1* c.2647T>C (p.Ter883Glnext*29) | --- | **PM4 is not applied to small in-frame indels because the impact of amino acid level changes of *CDH1* variants is inconclusive.** |
| PM5 | Novel missense change at amino acid residue where a different missense variant is pathogenic | --- | --- | --- | --- | **PM5_supporting is applicable to nonsense and frameshift variants that are predicted/proved to undergo NMD.**  **Site-specific recommendations for the application of PM5_Supporting for canonical splicing variants are provided in the splicing table.** | **The nonsense or frameshift variant must not impact splicing based on RNA assay or splicing predictions.** |
| PM6 | Assumed *de novo*, but w/o confirmation of paternity and maternity | --- | ≥Four patients **meet the HDGC individual phenotype criteria** w/o parental confirmation | ≥Two patients **meet the HDGC individual phenotype criteria** w/o parental confirmation | One patient **meet the HDGC individual phenotype criteria** w/o parental confirmation | --- | Use ClinGen’s *de novo* point system for a highly specific phenotype |
| PP1 | Cosegregation in multiple affected family members in a gene definitively known to cause the disease | --- | --- | ≥Seven **informative** meioses across ≥2 families | Five-six **informative** meioses across ≥1 famil**y** | Three-four **informative** meioses across ≥1 famil**y** | Based strength of rule code on number of meioses across one or more families |
| PP2 | Missense variant in a gene with a low rate of benign missense variation & where missense variants are a common mechanism of disease | --- | --- | --- | --- | --- | **Not applicable for *CDH1*** |
| PP3 | Multiple lines of computational evidence support a deleterious effect on the gene or gene product | --- | --- | --- | Variants affecting the same splice site as a well-characterized variant with similar or worse *in silico*/ RNA predictions | At least three *in silico* splicing predictors in agreement **(SpliceAI, MaxEntScan, SSF, GeneSplicer, HSF, TraP)** | **PP3 cannot be applied for canonical splice sites.** PP3 code also does not apply to the last nucleotide of exon 3 (c.387G). Do not use protein-based computational prediction models for missense variants. |
| PP4 | Patient’s phenotype or family history is highly specific for a disease with a single genetic etiology | --- | --- | --- | --- | --- | **Not applicable for *CDH1*** |
| PP5 | Reputable source recently reports variant as pathogenic | --- | --- | --- | --- | --- | **Not applicable for *CDH1*** |
| BA1 | Allele frequency is greater than expected for disorder | MAF cutoff of 0.2% | --- | --- | --- | --- | 99.99% CI; subpopulation must **≥ 2,000 alleles and** have a minimum of five **variant** alleles present |
| BS1 | Allele frequency is greater than expected for disorder | --- | MAF cutoff of 0.1% | --- | --- | --- | 99.99% CI; subpopulation must **≥ 2,000 alleles and** have a minimum of five **variant** alleles present.  **We allow a variant to reach a likely benign classification based on BS1 alone.** |
| BS2 | Observed in a healthy adult individual for a dominant disorder with full penetrance expected at an early age | --- | --- | Variant seen in ≥10 individuals w/o **GC,** DGC, **g**SRC tumors, or LBC & whose families do not suggest HDGC | --- | Variant seen in ≥3 individuals w/o **GC,** DGC, **g**SRC tumors, or LBC & whose families do not suggest HDGC | **We allow a variant to reach a likely benign classification based on BS2 alone.**  **BS2 cannot be applied to variants in which more than 30% of reported individuals meet HDGC criteria.** |
| BS3 | Well-established *in vitro* or *in vivo* functional studies show no damaging effect on protein function or splicing | --- | --- | Functional RNA studies demonstrating no impact on transcript composition | --- | --- | This rule can only be used to demonstrate lack of splicing **and can only be applied to Synonymous, Intronic or Non-coding variants.**  BS3 may be downgraded based on quality of data |
| BS4 | Lack of segregation in affected members of a family | --- | --- | Per original ACMG/AMP guidelines | --- | --- | Beware of the presence of phenocopies (e.g., breast cancer) that can mimic lack of segregation. Also, families may have more than one pathogenic variant contributing to another AD disorder |
| BP1 | Missense variant in a gene for which primarily truncating variants are known to cause disease | --- | --- | --- | --- | --- | **Not applicable for *CDH1*** |
| BP2 | Observed in a healthy homozygous individual, or in *trans* with a pathogenic variant for a fully penetrant dominant gene/disorder or observed in *cis* with a pathogenic variant | --- | --- | Variant observed *in trans* w/known pathogenic variant (phase confirmed) **OR** observed in the homozygous state in individual w/o personal &/or family history of DGC, LBC, or SRC tumors | --- | Variant is observed *in cis* (or phase is unknown) w/ a pathogenic variant  **OR observed in the homozygous state in gnomAD** | Evidence code is dependent on the strength of data. Take consideration of the quality of sequencing data when applying code. |
| BP3 | In-frame deletions/insertions in a repetitive region without a known function | --- | --- | --- | --- | --- | **Not applicable for *CDH1*** |
| BP4 | Multiple lines of computational evidence suggest no impact on gene/gene product | --- | --- | --- | --- | Splicing predictions only. At least three *in silico* splicing predictors in agreement **(SpliceAI, MaxEntScan, SSF, GeneSplicer, HSF, TraP)** | Do not use protein based computational prediction models **and BP4 is not applicable for missense variants.** |
| BP5 | Variant found in a case with an alternate molecular basis for disease | --- | --- | --- | --- | Per original ACMG/AMP guidelines | This applies if a P/LP variant is identified in an alternate gene known to cause HDGC (**currently only** *CTNNA1*) |
| BP6 | Reputable source recently reports variant as benign | --- | --- | --- | --- | --- | **Not applicable for *CDH1*** |
| BP7 | Synonymous variant which splicing prediction algorithms predict no impact to the splice consensus sequence nor the creation of a new splice site & the nucleotide is not highly conserved. | --- | --- | --- | --- | Synonymous **and intronic variants at or beyond +7 to -21 locations.** | **Note the *CDH1* rule specification does not require a conservation prediction.** We allow use of BP7 with BP4, as appropriate, to classify variants meeting both criteria as likely benign. |

**Table 2: Variants classified as Likely Benign based on BS2 alone**

| ***CDH1* Variants (NM_004360.5)** | **Applied Evidence Codes** | **No. of individuals without HDGC phenotypes** | **ClinVar Variation Id** | **Previous ClinVar Assertion** | **VCEP Assertion** |
| --- | --- | --- | --- | --- | --- |
| c.4G>A (p.Gly2Ser) | BS2 | 20 | 183998 | Uncertain | Likely Benign |
| c.8C>G (p.Pro3Arg) | BS2 | 73 | 142469 | Conflicting | Likely Benign |
| c.32TGC[7] (p.Leu14_Leu15dup) | BS2, PM2_Supporting | 21 | 142455 | Uncertain | Likely Benign |
| c.32TGC[6] (p.Leu15dup) | BS2 | 25 | 220798 | Uncertain | Likely Benign |
| c.113C>A (p.Thr38Lys) | BS2 | 10 | 231383 | Uncertain | Likely Benign |
| c.113C>T (p.Thr38Met) | BS2, PM2_Supporting | 11 | 133850 | Uncertain | Likely Benign |
| c.163+4_163+6dup | BS2 | 14 | 186186 | Conflicting | Likely Benign |
| c.164T>G (p.Val55Gly) | BS2 | 113 | 132769 | Conflicting | Likely Benign |
| c.269G>A (p.Arg90Gln) | BS2 | 55 | 142692 | Conflicting | Likely Benign |
| c.499G>A (p.Glu167Lys) | BS2 | 28 | 219431 | Uncertain | Likely Benign |
| c.670C>T (p.Arg224Cys) | BS2 | 392 | 127932 | Uncertain | Likely Benign |
| c.866C>T (p.Ala289Val) | BS2 | 12 | 216597 | Uncertain | Likely Benign |
| c.1118C>T (p.Pro373Leu) | BS2 | 38 | 142285 | Uncertain | Likely Benign |
| c.1225T>C (p.Trp409Arg) | BS2 | 40 | 133855 | Uncertain | Likely Benign |
| c.1297G>A (p.Asp433Asn) | BS2 | 110 | 127910 | Conflicting | Likely Benign |
| c.1493A>C (p.Asp498Ala) | BS2, PM2_Supporting | 10 | 234528 | Uncertain | Likely Benign |
| c.1865A>G (p.Asn622Ser) | BS2 | 25 | 136062 | Uncertain | Likely Benign |
| c.2053G>A (p.Val685Met) | BS2 | 12 | 133847 | Uncertain | Likely Benign |
| c.2076_2077inv (p.Gly693Ser) | BS2 | 41 | 220300 | Uncertain | Likely Benign |
| c.2131C>G (p.Leu711Val) | BS2, PM2_Supporting | 32 | 12231 | Uncertain | Likely Benign |
| c.2329G>A (p.Asp777Asn) | BS2 | 248 | 127922 | Conflicting | Likely Benign |
| c.2343A>T (p.Glu781Asp) | BS2 | 189 | 127923 | Conflicting | Likely Benign |
| c.2512A>G (p.Ser838Gly) | BS2 | 188 | 12233 | Conflicting | Likely Benign |
| c.2635G>A (p.Gly879Ser) | BS2 | 289 | 127928 | Conflicting | Likely Benign |

**Table S1: 2020 hereditary diffuse gastric cancer (HDGC) genetic testing criteria**

| **Family criteria, where family members are first-degree or second-degree blood relatives of each other:** | |
| --- | --- |
| **1** | **≥2 cases of gastric cancer in family regardless of age, with at least one DGC** |
| **2** | **≥1 case of DGC at any age in family, and ≥1 case of LBC at age <70 years, in different family members** |
| **3** | **≥2 cases of LBC in family members <50 years of age** |
| **Individual criteria:** | |
| **4** | **DGC at age <50 years** |
| **5** | **DGC at any age in individuals of Maori ethnicity** |
| **6** | **DGC at any age in individuals with a personal or family history (first-degree relative) of cleft lip or cleft palate** |
| **7** | **History of DGC and LBC, both diagnosed <70 years** |
| **8** | **Bilateral LBC, diagnosed at age <70 years** |
| **9** | **Gastric *in situ* signet ring cells (SRC) or pagetoid spread of SRCs in individuals <50 years of age** |

**Table S2: Curations of five initiation codon variants**

|  |  | **Version 1 criteria codes applied, where PVS1_Moderate was applied for initiation codon variations** | | **Version 2/3 criteria codes applied, where PVS1 was applied for initiation codon variations** | |
| --- | --- | --- | --- | --- | --- |
| ***CDH1* Variants**  **(NM_004360.5)** | **No. of probands or families meeting HDGC criteria** | **Criteria codes applied**  **(PVS1_Moderate)** | **Variant classification (PVS1_Moderate)** | **Criteria codes applied**  **(PVS1)** | **Variant classification**  **(PVS1)** |
| **c.1A>G (p.Met1?)** | **1** | **PVS1_Moderate, PM2_Supporting, PP1,**  **PS4_Supporting** | **Uncertain Significance** | **PVS1, PM2_Supporting, PP1, PS4_Supporting** | **Pathogenic** |
| **c.2T>G (p.Met1?)** | **2** | **PVS1_Moderate, PM2_Supporting, PS4_Moderate** | **Uncertain Significance** | **PVS1, PM2_Supporting, PS4_Moderate** | **Pathogenic** |
| **c.2T>A (p.Met1?)** | **1** | **PVS1_Moderate, PM2_Supporting, PS4_Supporting** | **Uncertain Significance** | **PVS1, PM2_Supporting, PS4_Supporting** | **Pathogenic** |
| **c.2T>C (p.Met1?)** | **2** | **PVS1_Moderate, PS4_Moderate, PM2_Supporting** | **Uncertain Significance** | **PVS1, PS4_Moderate, PM2_Supporting** | **Pathogenic** |
| **c.3G>A (p.Met1?)** | **4** | **PVS1_Moderate, PS4, PM2_Supporting, PP1_Moderate** | **Likely Pathogenic** | **PVS1,**  **PS4, PM2_Supporting, PP1_Moderate** | **Pathogenic** |

**Table S3: Splicing Table**

| **Exon** | **GT-AG 1,2 Splice site** | **Location** | **Prediction if exon skipping** | **RNA assay** | **Canonical splice site variants curated** | **Recommended PVS1 strength and the application of PM5_Supporting criterion** |
| --- | --- | --- | --- | --- | --- | --- |
| **Exon 1** | **Donor** | **c.48** | **Likely cryptic site** |  | **LP: c.48+1G>A** | **PVS1_Strong+PM5_Supporting** |
| **Exon 2** | **Acceptor** | **c.49** | **frameshift** | **frameshift** | **P: c.49-2A>G*** | **PVS1_Strong+PM5_Supporting** |
|  | **Donor** | **c.163** |  |  |  | **PVS1_Strong** |
| **Exon 3** | **Acceptor** | **c.164** | **frameshift** |  |  | **PVS1_Strong** |
|  | **Donor** | **c.387** |  | **in-frame transcripts** | **VUS: c.387+1G>A*** | **PVS1_Moderate** |
| **Exon 4** | **Acceptor** | **c.388** | **In-frame deletion (aa130-177)** |  |  | **PVS1_Strong** |
|  | **Donor** | **c.531** |  |  | **LP: c.531+1G>A** | **PVS1_Strong+PM5_Supporting** |
| **Exon 5** | **Acceptor** | **c.532** | **In-frame deletion (aa178-229)** |  | **LP: c.532-1G>C** | **PVS1_Strong+PM5_Supporting** |
|  | **Donor** | **c.687** |  |  | **LP: c.687+1G>A; c.687+2T>C** | **PVS1_Strong+PM5_Supporting** |
| **Exon 6** | **Acceptor** | **c.688** | **frameshift** |  |  | **PVS1_Strong** |
|  | **Donor** | **c.832** |  |  | **P: c.832+1G>T**  **LP: c.832+1G>A** | **PVS1_Strong+PM5_Supporting** |
| **Exon 7** | **Acceptor** | **c.833** | **frameshift** | **frameshift** | **P: c.833-2A>G*** | **PVS1_Strong+PM5_Supporting** |
|  | **Donor** | **c.1008** |  | **frameshift with cryptic site** | **LP: c.1008+2T>C; c.1008G>A*; c.1008G>T*** | **PVS1_Strong+PM5_Supporting** |
| **Exon 8** | **Acceptor** | **c.1009** | **In-frame deletion (aa 337-379)** |  |  | **PVS1_Strong** |
|  | **Donor** | **c.1137** |  | **frameshift with**  **cryptic site** | **P: c.1137+1delG; c.1137G>A***  **LP: c.1137+1G>A; c.1137+2T>C** | **PVS1_Strong+PM5_Supporting** |
| **Exon 9** | **Acceptor** | **c.1138** | **In-frame deletion (aa 380-440)** |  |  | **PVS1_Strong** |
|  | **Donor** | **c.1320** |  | **in-frame deletion (exon 9 skipping)** | **LP: c.1320+1G>C*** | **PVS1_Strong+PM5_Supporting** |
| **Exon 10** | **Acceptor** | **c.1321** | **frameshift** |  |  | **PVS1_Strong** |
|  | **Donor** | **c.1565** |  |  | **P: c.1565+1G>C; c.1565+1G>A;**  **c.1565+1G>T; c.1565+2dupT**  **LP: c.1565+1delG** | **PVS1_Strong+PM5_Supporting** |
| **Exon 11** | **Acceptor** | **c.1566** | **frameshift** | **predicted in-frame insertion with potential rescue transcript** | **VUS: c.1566-1G>C; c.1566-2A>G** | **PVS1_Moderate** |
|  | **Donor** | **c.1711** |  |  | **LP: c.1711+1G>C; c.1711+1G>A;**  **c.1711+2_1711+7delTAAGGG** | **PVS1_Strong+PM5_Supporting** |
| **Exon 12** | **Acceptor** | **c.1712** | **In-frame deletion (aa 571-646)** | **in-frame deletion (c.1712_1720del9)** | **VUS: c.1712-2A>C*** | **PVS1_Moderate** |
|  | **Donor** | **c.1936** |  |  |  | **PVS1_Strong** |
| **Exon 13** | **Acceptor** | **c.1937** | **In-frame deletion (aa 646-722)** |  |  | **PVS1_Strong** |
|  | **Donor** | **c.2164** |  |  | **VUS: c.2164+2T>C**  **VUS: c.2164+2dup (this variant actually affects +3 location)** | **PVS1_Strong** |
| **Exon 14** | **Acceptor** | **c.2165** | **frameshift** |  | **LP: c.2165-1G>C** | **PVS1_Strong+PM5_Supporting** |
|  | **Donor** | **c.2295** |  |  |  | **PVS1_Strong** |
| **Exon 15** | **Acceptor** | **c.2296** | **In-frame deletion (aa 766-813)** |  | **LP: c.2296-1G>A; c.2296-2A>G** | **PVS1_Strong+PM5_Supporting** |
|  | **Donor** | **c.2439** |  |  |  | **PVS1_Strong** |
| **Exon 16** | **Acceptor** | **c.2440** | **Likely cryptic site** | **abnormal splicing** | **LP: c.2440-2A>G*** | **PVS1_Strong+PM5_Supporting** |

*** RNA functional assay performed.**

**Table S4: 5 Pathogenic/Likely Pathogenic predicted missense substitution variants**

| **Missense Variants**  **NM_004360.5(*CDH1*)** | **RNA splicing assay** | **Criteria codes applied** | **Classification** |
| --- | --- | --- | --- |
| **c.715G>A (p.Gly239Arg)** | **Abnormal out of frame transcript with the first 29 base pairs of exon 6 deleted (PMID: 17545690)** | **PS4_Very Strong, PS3, PM2_Supporting, PP3** | **Pathogenic** |
| **c.1008G>T (p.Glu336Asp)** | **Cryptic splice site with 7bp intron inclusion and early PTC in exon 8 (PMID: 9537325)** | **PS3, PM2_Supporting, PVS1_Moderate, PS4_Supporting** | **Likely pathogenic** |
| **c.1057G>A (p.Glu353Lys)** | **Abnormal out of frame splicing with r.1055_1137del (PMID: 32133419)** | **PS3, PS4_Moderate, PM2_Supporting, PP3** | **Likely pathogenic** |
| **c.1679C>G (p.Thr560Arg)** | **Created a novel 5‘ splice donor site with a 32 bp deletion in exon 11 (PMID: 27880784)** | **PS3, PS4, PP1_Strong, PM2_Supporting, PP3** | **Pathogenic** |
| **c.2195G>A (p.Arg732Gln)** | **Complex splicing and deletion of 32 base pairs at the start of exon 14 (PMID: 17545690)** | **PS3, PS4_Moderate, PM2_Supporting, PP3** | **Likely pathogenic** |

**Table S5: Application of PM5_supporting to nonsense/frameshift variants that are predicted/proved to undergo NMD**

| **Exon** | **Location** | **No. of nonsense/frameshift curated (N=113) before applying PM5_Supporting** |
| --- | --- | --- |
| **Exon 1** | **c.1-c.48** | **P (1): c.26C>A (p.Ser9Ter)**  **LP (2): c.11G>A (p.Trp4Ter); c.12G>A (p.Trp4Ter)** |
| **Exon 2** | **c.49-c.163** | **P (4): c.59G>A (p.Trp20Ter); c.60G>A (p.Trp20Ter); c.70G>T (p.Glu24Ter); c.124_126delCCCinsT (p.Pro42Serfs)**  **LP (1): c.76G>T (p.Glu26Ter)** |
| **Exon 3** | **c.164-c.387** | **P (8): c.187C>T (p.Arg63Ter); c.208dup (p.Ser70Phefs); c.220C>T (p.Arg74Ter); c.283C>T (p.Gln95Ter); c.308G>A**  **(p.Trp103Ter); c.360dup (p.His121fs); c.377del (p.Pro126Argfs); c.382delC (p.His128Ilefs)**  **LP (5): c.202delT (p.Tyr68Ilefs); c.261delG (p.Arg87Serfs); c.315delC (p.Thr106Profs); c.337A>T (p.Lys113Ter);**  **c.369_375CCGCCCC[3] (p.His128fs)** |
| **Exon 4** | **c.388-c.531** | **P (6): c.454_460delCAGAAGA (p.Gln152Glufs); c.480_486delinsAGAATA (p.Ile161fs); c.489C>A (p.Cys163Ter);**  **c.521dupA (p.Asn174Lysfs); c.504delA (p.Gly169Alafs); c.529C>T (p.Gln177Ter)**  **LP (7): c.436_437TC[1] (p.Pro147fs); c.454_460dup (p.Arg154Thrfs); c.455_465delAGAAGAGAGAC (p.Gln152Leufs);**  **c.457_460delAAGA (p.Lys153Glufs); c.457A>T (p.Lys153Ter); c.467G>A (p.Trp156Ter); c.468G>A (p.Trp156Ter)** |
| **Exon 5** | **c.532-c.687** | **P (2): c.603delT (p.Val202Leufs); c.656del (p.Pro219fs)**  **LP (1): c.594_595insT (p.Thr199fs)** |
| **Exon 6** | **c.688-c.832** | **P (1): c.720del (p.Asn240fs)**  **LP (4): c.692_693TC[2] (p.His233fs); c.707C>A (p.Ser236Ter); c.781G>T (p.Glu261Ter); c.793G>T (p.Glu265Ter)** |
| **Exon 7** | **c.833-c.1008** | **P (2): c.940A>T (p.Lys314Ter); c.1003C>T (p.Arg335Ter)** |
| **Exon 8** | **c.1009-c.1137** | **P (7): c.1009_1010delAG (p.Ser337Phefs); c.1023T>G (p.Tyr341Ter); c.1051C>T (p.Gln351Ter); c.1064delT (p.Leu355Terfs);**  **c.1085delT (p.Val362Glyfs); c.1107del (p.Asn369Lysfs); c.1131del (p.Thr378fs)**  **LP (1): c.1031_1032dup (p.Val345fs)** |
| **Exon 9** | **c.1138-c.1320** | **P (2): c.1147C>T (p.Gln383Ter); c.1235_1236TA[3] (p.Ile415fs)**  **LP (2): c.1170del (p.Asn390fs): c.1312del (p.Thr438fs)** |
| **Exon 10** | **c.1321-c.1565** | **P (4): c.1408del (p.Thr470fs); c.1476_1477delAG (p.Arg492Serfs); c.1488_1494delCGAGGAC (p.Glu497Leufs);**  **c.1531C>T (p.Gln511Ter)**  **LP (7): c.1341del (p.Lys447fs); c.1354_1357del (p.Leu452fs); c.1390del (p.Val464fs); c.1443del (p.Asn481fs);**  **c.1460_1461del (p.Val487Alafs); c.1480G>T (p.Glu494Ter); c.1505delG (p.Gly502Alafs)** |
| **Exon 11** | **c.1566-c.1711** | **P (4): c.1578G>A (p.Trp526Ter); c.1587dup (p.Ala530fs); c.1590dup (p.Asn531fs); c.1612delG (p.Asp538Thrfs)**  **LP (3): c.1569T>A (p.Tyr523Ter); c.1636delG (p.Ala546Leufs); c.1679dup (p.Tyr561Valfs)** |
| **Exon 12** | **c.1712-c.1936** | **P (6): c.1733dup (p.Gly579fs); c.1779dup (p.Ile594fs); c.1792C>T (p.Arg598Ter); c.1895_1896delAC (p.His632Argfs);**  **c.1913G>A (p.Trp638Ter); c.1921C>T (p.Gln641Ter)**  **LP (2): c.1746dup (p.Leu583Alafs); c.1917_1918del (p.Ile640fs)** |
| **Exon 13** | **c.1937-c.2164** | **P (5): c.1979dup (p.Gly661_Asp662insTer); c.1999del (p.Leu667fs); c.2062_2063TG[1] (p.Cys688_Glu689delinsTer);**  **c.2095C>T (p.Gln699Ter); c.2100del (p.Val701Serfs);**  **LP (8): c.1942G>T (p.Glu648Ter); c.1948_1949del (p.Ile650HisfsTer12); c.1948_1949dup (p.Ile651Serfs);**  **c.1993del (p.Ile665Serfs); c.2029dup (p.Gln677Profs); c.2076_2077del (p.Gly693ArgfsTer3); c.2104G>T (p.Glu702Ter);**  **c.2144delG (p.Gly715Glufs)** |
| **Exon 14** | **c.2165-c.2295** | **P (4): c.2265T>A (p.Tyr755Ter); c.2276delG (p.Gly759Glufs); c.2287G>T (p.Glu763Ter); c.2293C>T (p.Gln765Ter)**  **LP (1): c.2272G>T (p.Glu758Ter)** |
| **Exon 15** | **c.2296-c.2439** | **P (2): c.2398delC (p.Arg800Alafs); c.2430delT (p.Phe810Leufs)**  **LP (3): c.2311C>T (p.Gln771Ter); c.2324delG (p.Gly775Alafs); c.2386dup (p.Arg796Profs)** |
| **Exon 16** | **c.2440-c.2649** | **P (1): c.2506G>T (p.Glu836Ter)**  **LP (2): c.2446A>T (p.Lys816Ter); c.2490dup (p.Leu831Alafs)**  **VUS (5): c.2505_2506dup (p.Glu836fs); c.2526delT, p.(Ala843Leufs); c.2547_2548insA (p.Ser850fs);**  **c.2549_2550delCC (p.Ser850Phefs); c.2594G>A (p.Trp865Ter)** |
